# Public Covid-19 X-ray datasets and their impact on model bias - a systematic review of a significant problem

**DOI:** 10.1101/2021.02.15.21251775

**Authors:** Beatriz Garcia Santa Cruz, Matías Nicolás Bossa, Jan Sölter, Andreas Dominik Husch

**Affiliations:** Centre Hospitalier de Luxembourg, 4, Rue Ernest Barble, L-1210 Luxembourg, Luxembourg; Luxembourg Centre for Systems Biomedicine, University of Luxembourg, 7, Avenue des Hauts Fourneaux, L-4362 Esch-sur-Alzette, Luxembourg

**Keywords:** COVID-19, machine learning, datasets, X-Ray, imaging, review, bias, confounding

## Abstract

Computer-aided-diagnosis for COVID-19 based on chest X-ray suffers from weak bias assessment and limited quality-control. Undetected bias induced by inappropriate use of datasets, and improper consideration of confounders prevents the translation of prediction models into clinical practice. This study provides a systematic evaluation of publicly available COVID-19 chest X-ray datasets, determining their potential use and evaluating potential sources of bias.

Only 5 out of 256 identified datasets met at least the criteria for proper assessment of risk of bias and could be analysed in detail. Remarkably almost all of the datasets utilised in 78 papers published in peer-reviewed journals, are not among these 5 datasets, thus leading to models with high risk of bias. This raises concerns about the suitability of such models for clinical use.

This systematic review highlights the limited description of datasets employed for modelling and aids researchers to select the most suitable datasets for their task.

## 1. Introduction

Less than a year has passed since the novel corona virus SARS-CoV-2 gained world wide attention and eventually developed to the global COVID-19 pandemic (Sohrabi et al. (2020)). Diagnosis plays a vital role in the management of cases and the allocation of potentially limited resources, like hospital/ICU beds. Hence, there is an urgent necessity to create trustworthy tools for diagnosis and prognosis of the disease. While most of the people with COVID-19 infection do not develop pneumonia (Cleverley et al. (2020)), the early identification of COVID-19 induced pneumonia cases is essential. To this end, imaging studies such as planar X-ray and computed tomography (CT) are employed. Chest X-ray (CXR) is a widely available, fast, non-invasive, and relatively cheap tool to diagnose and monitor COVID-19 induced pneumonia (Aljondi and Alghamdi (2020)). In contrast to CT, CXR yields lower diagnostic sensitivity, however this is outweighed by the ease of application, even in patients in intensive care treatment using portable scanners.

Currently, no distinct feature specific for COVID-19 pneumonia is known in chest imaging. However, a combination of multi-focal peripheral lung changes, mainly ground glass opacities and/or consolidations, often in a bilateral arrangement, has been demonstrated (Cleverley et al.(2020)).

### 1.1. Motivation

Machine learning, and in particular deep learning methods, promise to assist medical staff in coherent diagnosis and interpretation of images (Choy et al. (2018); McBee et al. (2018)). A remarkable amount of machine learning models has been proposed in a very short amount of time to tackle the problems of COVID-19 diagnosis, quantification and prognosis from X-Ray imaging (Shoeibi et al. (2020); Islam et al. (2020); Ilyas et al. (2020)).

However, there is growing awareness in the community that the presence of different sources of bias significantly decreases the overall generalisation ability of the models, leading to overestimated model performance reported in internal validation compared to evaluation on independent test data (Soneson et al. (2014); Cohen et al. (2020b); Zech et al. (2018); Maguolo and Nanni (2020)). In addition, numerous journal editorials are calling for better development, evaluation and reporting practices of machine learning models aimed for clinical application (Mateen et al. (2020); Nagendran et al. (2020); Campbell et al. (2020); The Lancet Digital Health (2020); O’Reilly-Shah et al. (2020); The Lancet Digital Health (2019); Stevens et al. (2020)). Underneath these caveats are the growing concerns about ethics and risk of harmful outcomes of using AI in medical applications (Campolo et al. (2018); Geis et al. (2019); Brady and Neri (2020)).

In order to avoid or at least be able to detect potential bias, it is important that datasets and models are well documented. Some aspects of dataset building, such as criteria for subject inclusion and exclusion, recruitment method, patterns in missing data, and many more, may influence model accuracy and introduce bias in prediction models. Among the most common sources of bias are unknown confounders and selection bias. (Steyerberg (2009); Griffith et al. (2020); Greenland et al. (1999); Heckman (1979)). In both cases, the presence of spurious association between predictors and outcomes might be learned by the model, leading to undetected overfitting resulting in models not capable of generalizing and eventually failing in clinical application (Wolff et al. (2019), 1.1).

#### 1.1.1. State-of-the-Art

In a recent review and critical appraisal of prediction models for diagnosis and prognosis of COVID-19 (Wynants et al. (2020)) *all* evaluated models were rated at high risk of bias. The authors arrived to the conclusion that they “do not recommend any of these reported prediction models for use in current practice”. For the subset of diagnostic models based on medical imaging two main causes for high risk of bias were identified: 1) a lack of information to assess selection bias (such as how controls were selected or which subset of patients underwent imaging) and 2) a lack of clear reporting of image annotation procedures and quality control measures. Similar conclusions were obtained in another publication specifically addressing machine learning models using chest X-ray and CT images (Roberts et al. (2021)). They found high or unclear risk of bias in *all* studies and that the reported results were extremely optimistic, mainly due to limitation in the datasets or combination of datasets used.

### 1.2. Aim & Hypothesis

Given the previously described disappointing state-of-the-art, we hypothesise that the current main obstacle towards building clinically applicable machine learning models for COVID-19 is *not* the machine learning techniques *per se*, but instead access to reliable training data that on the one hand captures the problem complexity, but on the other does not induce undetected bias to the models.

Therefore, there is a need to raise awareness of such problems and to aid modellers to efficiently find the right dataset for their particular problem supporting efficient creating of robust models. This paper gives an overview on current publicly available chest X-ray datasets, identifying strengths as well as limitations, including most evident potential sources of bias.

### 1.3. Paper structure

We systematically evaluated the quality of COVID-19 chest X-ray datasets and their utility for training prediction models using an adapted version of the CHARMS tool (Critical Appraisal and Data Extraction for Systematic Reviews of Prediction Modelling Studies, Moons et al. (2014)). Dataset quality is measured by the amount and the detail in the description of the dataset variables and of the dataset building process. Model designers needs this information to evaluate the risk of bias and the generalizability, for example, using tools such as PROBAST (Prediction model Risk Of Bias ASsessment Tool, Wolff et al. (2019)). The utility is determined by the structure and amount of information, for example, only those datasets including information about patient survival can be used for training prognosis models of risk of death. Thus this paper provides a more in depth description of datasets than previous works (Shuja et al. (2020); Sohan (2020); Shoeibi et al. (2020); Islam et al. (2020); Ilyas et al. (2020)), that have focused on surveying papers describing methods, and on identifying the datasets used to train these methods, without assessing the datasets quality or utility.

In Section 2 the methodology used to systematic search for datasets and evaluate their potential for clinical prediction models is given. Next, in Section 3, the datasets selected for review are analysed, describing their information content. A general overview of some of the datasets most commonly used in published paper is also presented in order to put this review in the context of the current model development scenario. The landscape of interrelated datasets is complex. The most frequently used datasets are compositions of many individual sources and thus also the ones that are most difficult to assess for the risk of bias. In Section 4 the insight extracted from this analysis is commented. Finally, some recommendations for researchers aiming at clinical prediction model building are given in Section 5.

## 2. Methods

In this section, the main tools to address prediction model quality (PROBAST, TRIPOD and TREE) are briefly introduced and most relevant aspects concerning datasets are highlighted. Subsequently, the search strategy (based on PRISMA) adopted to find datasets with available information required to assess quality is presented. The inclusion and exclusion criteria are defined according to the prediction model quality guidelines. Next, a data extraction tool (CHARMS) is adapted to evaluate eligible datasets, verifying that all relevant information needed for assessing model quality is present. Finally, and also following PRISMA recommendations, published paper presenting Machine Learning models using COVID-19 X-ray images were analysed to extract dataset usage statistics.

### 2.1. Tools for model evaluation

There are several tools to evaluate prediction model quality, risk of bias and transparency, depending on model characteristics. The elements of these tools specifically addressing aspects of the *data* are considered in this work.

PROBAST tool (Wolff et al. (2019)) was developed to evaluate the risk of bias and the applicability to the intended population and setting of diagnostic and prognostic prediction model studies. It requires to answer specific questions about participant selection criteria and setting, its numbers, information about predictors and outcomes, and whether all of these choices were appropriate for the model intended use. Whenever this appropriateness can not be answered, for example, because some aspect of the datasets is not properly or sufficiently described, the model is rated as having a hsigh risk of bias.

TRIPOD (Transparent Reporting of a Multivariable Prediction Model for Individual Prognosis or Diagnosis, Moons et al. (2015)) aims to improve reporting and understanding of prediction model studies. Similarly to the PROBAST tool, it requires a clear definition of predictors and outcomes and a description of participant eligibility criteria, plus many more items.

A third example of quality assessment tool, in this case specifically designed for machine learning and artificial intelligence research, is given by a set of 20 critical questions proposed in Vollmer et al. (2020), to account for transparency, reproducibility, ethics, and effectiveness (TREE). Among the questions is the following: “Are the data suitable to answer the clinical question—that is, do they capture the relevant real world heterogeneity, and are they of sufficient detail and quality?”. To answer this question it is required to evaluate data quality and detail, including “accuracy of data collection methods, sampling of participants, eligibility criteria, and missing data”.

### 2.2. Dataset search, eligibility & selection

The search strategy for publicly available COVID-19 chest X-ray datasets consisted of two complementary approaches: a direct search for datasets using a dataset search engine, known compilation websites as well as direct extraction of data sources from reviews, and an indirect search for papers that describe prediction models for COVID-19. The papers from the indirect search were subsequently analysed to extract the employed datasets yielding additional dataset records. PRISMA (Moher et al. (2015)) (Preferred Reporting Items for Systematic Reviews and Meta-Analyses) statement was adapted to this special situation, where the final object of interest are datasets instead of studies.

Many datasets provided online are combinations of *primary datasets*, or even super-aggregations of other compilations. Moreover, the employ of such compilation datasets is very widespread in COVID-19 X-ray models. The aggregated datasets have complex inter-dependencies (Garcia Santa Cruz et al. (2020)) that can be dangerous (DeGrave et al. (2020)) when the users do not consider the origin of the data and the potential limitations. For example, some compilations merged COVID-19 positive cases from *adults* with healthy controls from *children*, which imposes a high risk of fitting to this confounder when not addressed in the modelling (Garcia Santa Cruz et al. (2020)). In addition, there is little or no benefit over adding each of the needed primary datasets individually by the researchers building the model.

Therefore, all compilation datasets, including all datasets from *Kaggle*, were discarded from the detailed analysis for potential bias. Instead the primary datasets included in the compilations were analysed for eligibility in a later step individually. For completeness, an overview of the most frequently used compilation, primary datasets, and their relations, is presented in Fig. 3.

For the identification step of the direct search entries the *google research dataset search*^1^ was employed using the query “COVID-19” & “X-ray” & “dataset”. Additionally four compilation website were examined ^2 3 4^. Finally, review papers on the topic (Shuja et al. (2020); Ilyas et al. (2020); Islam et al. (2020); Shoeibi et al. (2020); Wynants et al. (2020); Pham et al. (2020); Roberts et al. (2021); Garcia Santa Cruz et al. (2020)) were screened for dataset candidates. For the indirect search, PubMed and preprint services (medRxiv, bioRxiv and arXiv) were queried with the search terms “COVID-19” & “X-ray” & “dataset”. All queries were restricted to the time interval between 1st of January 2020 to 1st of October 2020.

During the screening step of the direct search for datasets, all the entries that were duplicates, the aggregated datasets and the non-open datasets were removed. In the parallel indirect search to yield datasets from papers, all papers with less than 10 citations based on google scholar where filtered out and the remaining papers were analysed to extract the datasets employed. As in the direct search, datasets that were non-open, non-COVID-19, or not based on chest X-ray were discarded. Next, all datasets from the direct and indirect search were merged, and those that were *not sufficiently documented* either in a paper or project website (e.g. GitHub repositories) to assess potential bias where discarded.

Final eligibility for inclusion was limited to datasets that include COVID-19 cases and that were collected under the same protocol. Hence, datasets including only controls and collections of case reports were discarded. Such information is essential to assess model applicability, generalizability and to determine the risk of bias, according to PROBAST, TRIPOD or TREE guidelines.

### 2.3. Dataset information extraction

CHARMS checklist (Moons et al. (2014)) was designed as a data extraction tool for systematic review of prediction modelling studies, including machine learning models. It is not specifically designed to evaluate datasets, however a large part of its items are devoted to extract information from data used in the studies. Up to our knowledge, there are no other tools, protocols or statements, exclusively designed for dataset evaluation. Therefore, a simplified version of CHARMS checklist is proposed in this work (Supplementary material, Appendix 1), where sections regarding model characteristics (model type, evaluation metrics, performance and results) were discarded. Specifically, the following domains were kept: Data source, Participant description, Outcomes, Predictors, Sample size and Missing data. The domains about model (development, performance and evaluation), Results and interpretation were omitted.

*Participant* description, including recruitment method, inclusion and exclusion criteria, are needed to determine the applicability and generalizability of the model, whether the study population is representative of the target population, and to discard the presence of selection mechanisms that can introduce bias. Information about received treatments could be relevant if they affect the outcome of prognostic models.

*Outcomes* to be predicted will depend on the purpose of the model, i.e. whether it is a prognostic or a diagnostic models. Radiological findings, lesion segmentation and disease differential diagnosis could be suitable outcomes for diagnostic models, when they are measured close in time to the image acquisition. When there are multiple images from the same subject acquired at different time-points, a prognostic model could be trained, where the diagnosis from the later image is predicted from the earlier one. Time to death or discharge, or whether ICU, supplementary oxygen or other life support treatments were needed, can be used also in prognostic models.

Although outcomes based on image findings could sometimes be determined a posteriori (e.g. with a post-hoc annotation by a radiologist), a precise definition of them is essential to describe model applicability. It is also important to specify whether outcomes where obtained blinded to predictors and/or the other way round, because this affects the causal model assumption and the strategies to mitigate bias (Castro et al. (2020)).

The main *predictor* of any model considered here is the X-ray lung image, however other measurements could be also used in the model. Details of the image acquisition protocol and acquisition device description are important as they could be a significant source of confounding when merging images from different sources. In addition, the model performance could be reduced if applied to images acquired using a different setting than in the training set.

Finally, a large enough *sample size* and the amount and treatment of *missing data* are highly relevant to avoid overfitting and confounding, respectively.

The full list of items used in this work are in Table S1 and detailed explanation of each item can be found in Moons et al. (2014).

### 2.4. Current landscape of COVID-19 X-ray datasets

As only few datasets met appropriate conditions for our final analysis, and for the sake of completeness, a collection of the most frequently used COVID-19 and non-COVID-19 datasets were included. Non-COVID-19 datasets are commonly employed to both, pre-train networks (Cohen et al. (2020a)), and as control cases in diagnosis models. A short description of each dataset was included as well as a diagram with their relationships. Additionally, we assessed the usage frequency of the selected datasets to depict the general trend.

### 2.5. Analysis of the dataset use frequency

Finally, in order to illustrate how little attention data quality has received by researchers so far, a temporal analysis of the dataset frequency use in published papers was conducted. During the identification step, published papers based on Machine Learning using publicly available datasets indexed in PudMed from May 2020 to December 2020 using the query (COVID-19, x-ray, deep learning) OR (COVID-19, x-ray, machine learning) OR (sars-cov-2,x-ray, deep learning) were included. Since we restricted our research to publications indexed on Pudmed, the record identification thought other sources as well as the removal step for duplication do not apply. Next, during the screening step, all the published manuscript which do not cover our initial research question, i.e. been an original research manuscripts using machine learning techniques X-ray modality, were removed. Subsequently, the datasets employed in each work were extracted. Those paper that not employed publicity available datasets or the datasets were not identifiable were removed during the eligibility step. Lastly, the temporal patters of dataset set use was analyse. PRISMA workflow of the process is depicted in supplementary material, Appendix 2.

## 3. Results

This sections consists of four parts. First, the results of the adapted PRISMA search are presented, next the eligible 5 datasets are analysed in detail. Afterwards, the most popular datasets, including the interactions of the aggregated datasets are described. Finally, the results of the frequency of datasets use are presented.

### 3.1. Dataset search & selection

During the direct search a total of 256 entries were identified; in particular, 175 from the google dataset search, 53 from compilations websites and 37 from review papers of COVID-19 datasets. In the indirect search a total of 415 papers were identified; specifically, 58 papers found in PubMed and the rest on pre-print services (195 in MedRviv, 93 in bioRxiv and 69 in arXiv).

The screening step of the direct search consisted on the removal of all duplicate entries (221) and non-open datasets (4). Duplicate datasets are remixes of previously published datasets and mainly found on kaggle (179). For the indirect research, pre-print with less that 10 publications (284) were filtered out, followed by 71 papers removed for the following reasons: duplicate papers (4), out-of-topic (42), not including any open dataset (20) and dataset information not accessible (5). Finally, a total of 60 papers that employed public datasets were kept and 31 public dataset were extracted. The adapted PRISMA protocol is represented in Fig. (2), where yellow boxes denote papers from the indirect search and yellow boxes denote datasets.

**Fig. 1.**
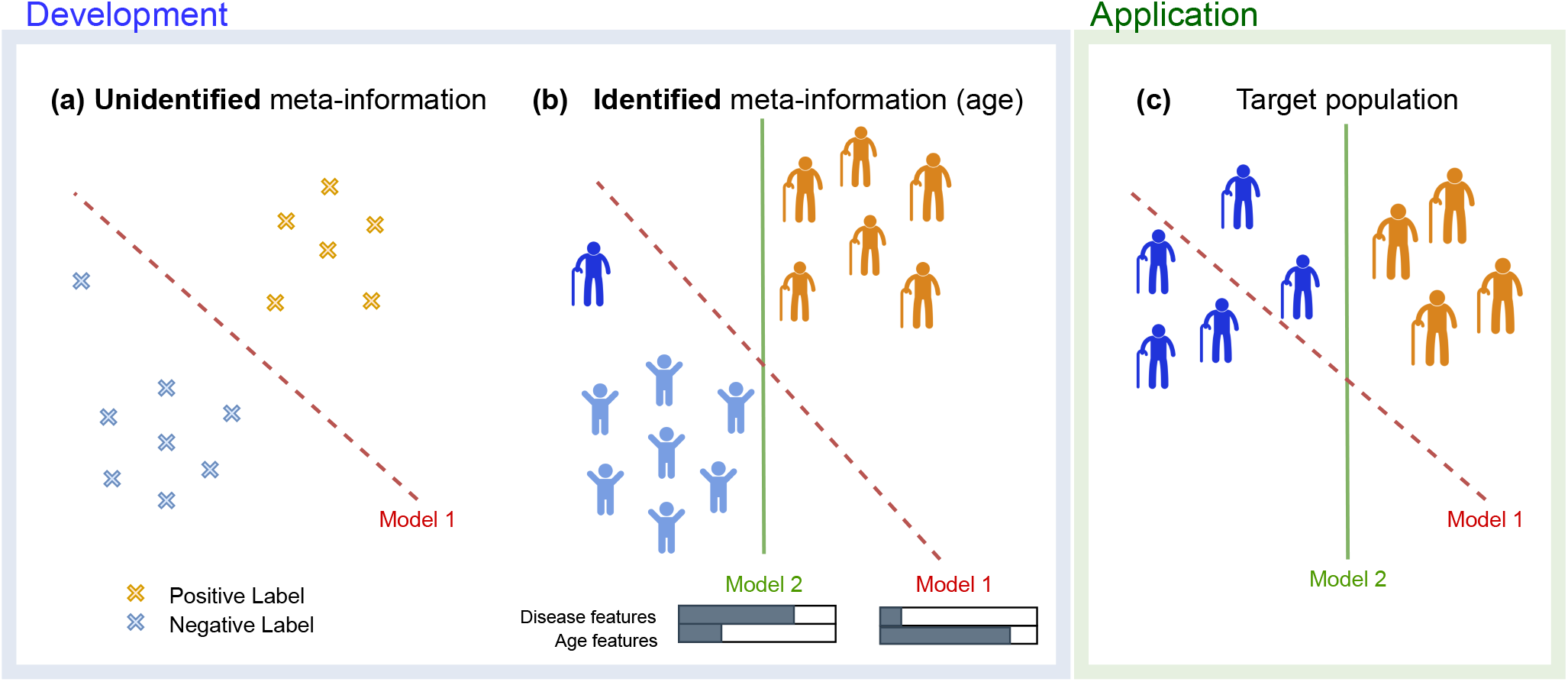
Importance of identified meta-information during model development. (a) Given a dataset with unidentified composition of the dataset population, there is a high risk of bias, i.e. a model is systematically prejudiced to faulty assumptions. (b) For example in an extreme case almost all of the control cases form a special sub-population of young age. With knowledge on the dataset age composition one is at least aware that any model developed with this dataset has a high risk of being biased by age (Model 1) or can even choose a model mitigating the age influence (Model 2). (c) Biased models are very likely to lead to impaired performance in the target population hampering generalizability.

**Fig. 2.**
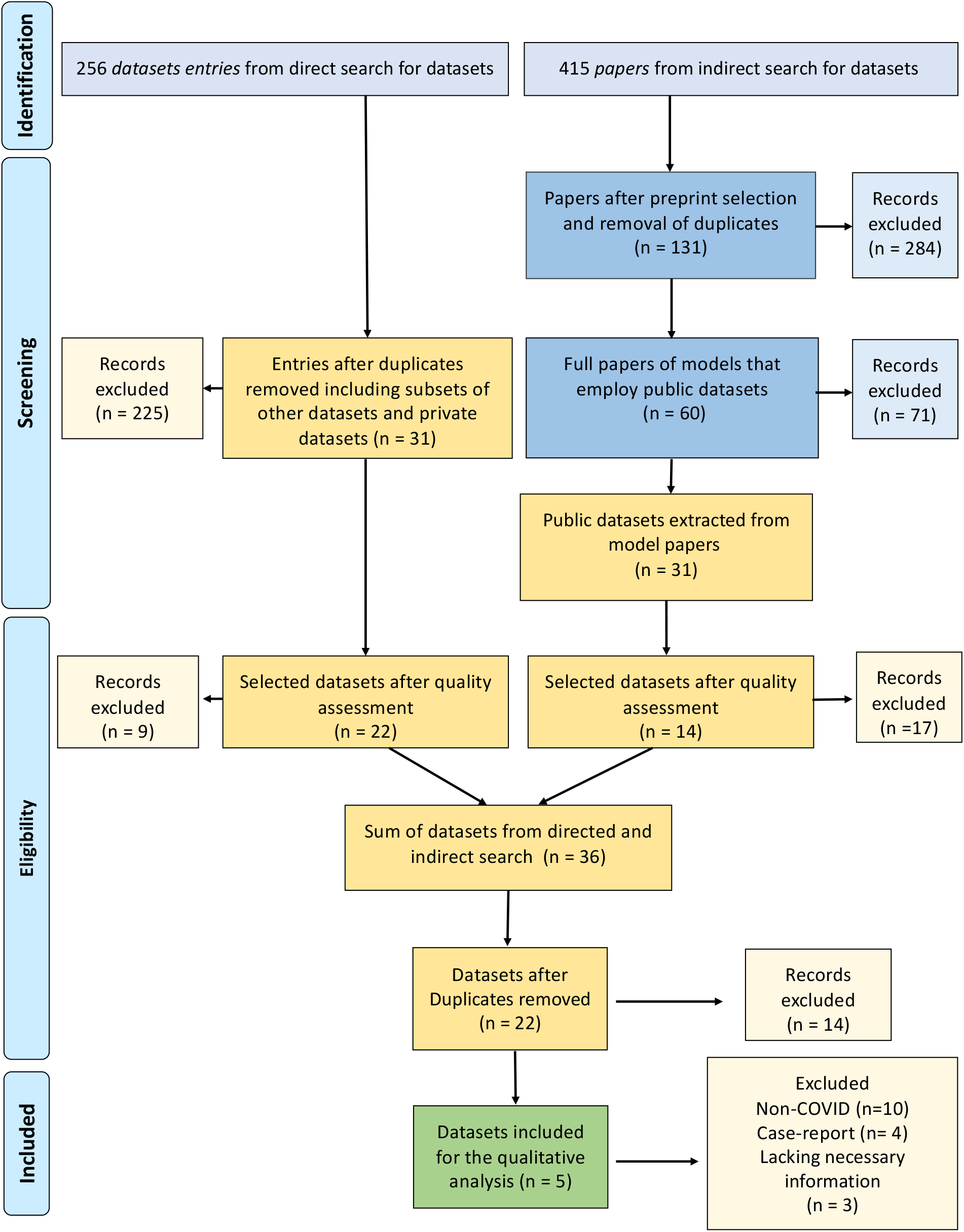
Adapted PRISMA workflow for the analysis of COVID-19 X-ray datasets. Boxes in blue indicate papers from the additional indirect search for papers *using* datasets, from which datasets (yellow/green) were extracted.

During the eligibility step, dataset not well documented from the direct (9) and indirect (16) search were removed. Next, all the selected dataset from both the direct and indirect strategy were merged and the duplicate removed.

Finally, in the inclusion step, datasets that do not contain COVID-19 cases were removed (10) as well as COVID-19 datasets that do not meet the inclusion criteria: case studies (4) or not collected under some prespecified protocol (3).

The selected 5 datasets included for the qualitative analysis were: *Brixia* dataset, from the ASST Spedali Civili (Civil Hospital) of Brescia, Italy; the *Cancer Imagine Archive* dataset from the University of Arkansas for Medical Sciences (USA), that is focused on underrepresented rural populations in the USA; *ML Hannover* (Winther et al. (2020)) from the Institute for Diagnostic and Interventional Radiology (Hannover, Germany); *HM Hospitals* dataset includes patients from the same Hospital Group in different cities in Spain and is focused on interactions between diagnosis, treatment and outcomes; and *BIMCV-COVID19* from the Medical Imaging Databank of the Valencia Region (<u>B</u>anco digital de <u>I</u>magen <u>M</u>edica de la <u>C</u>omunidad <u>V</u>alenciana). This last one comprises of differentsubsets: *BIM CV-COVID19*+ with two subsequent releases (iteration 1 and iteration 2), *BIMCV-COVID19-* and *BIMCV-COVID19-Padchest. BIMCV-COVID19*+ and *BIMCV-COVID19-* are collected from the same hospitals in the first half of 2020 and distinguished by either a positive or negative diagnostic test (PCR and/or Antigen). Because *BIMCV-COVID19-Padchest* contains cases collected at pre-COVID19 times and different hospitals, we do not include it for further analysis.

### 3.2. Dataset metadata availability for risk of biases assessment

For most of the datasets it was difficult or impossible to assess most of the information items. None of the datasets included an accompanying paper with a comprehensive description or study protocols. At the moment of writing this manuscript, *BIMCV* provided a description of the first iteration of the data (*BIMCV-COVID19*+ Vayá et al. (2020)) while further information, especially about the collection of the non-COVID-19 cases, is claimed to be coming soon. For the *Brixia* data, there is a detailed description of the radiological annotation process (Signoroni et al. (2020)).

#### Participants

There was limited information about participant eligibility and recruitment method, see Table 2 for details. All the datasets included participant sex, four of them included participant age, two of them included also subject height and weight, and only one included information about comorbidities, which is particular relevant in this disease because of the strong evidence of interactions between comorbidities and death risk (Yang et al. (2020)). In general, the participant description is too scarce to asses if the training population is representative of the target population, hampering the applicability of the models. It is also difficult to determine if there are selection mechanisms, including inclusion and exclusion criteria, that could be a source of strong confounding and limit generalizability and transportability of developed models. Only one dataset provides information about treatment, which is another serious limitation taking into account that this is a new disease and there are no prespecified treatment protocols and different experimental treatment were applied in each country or hospital unit.

**Table 1.**
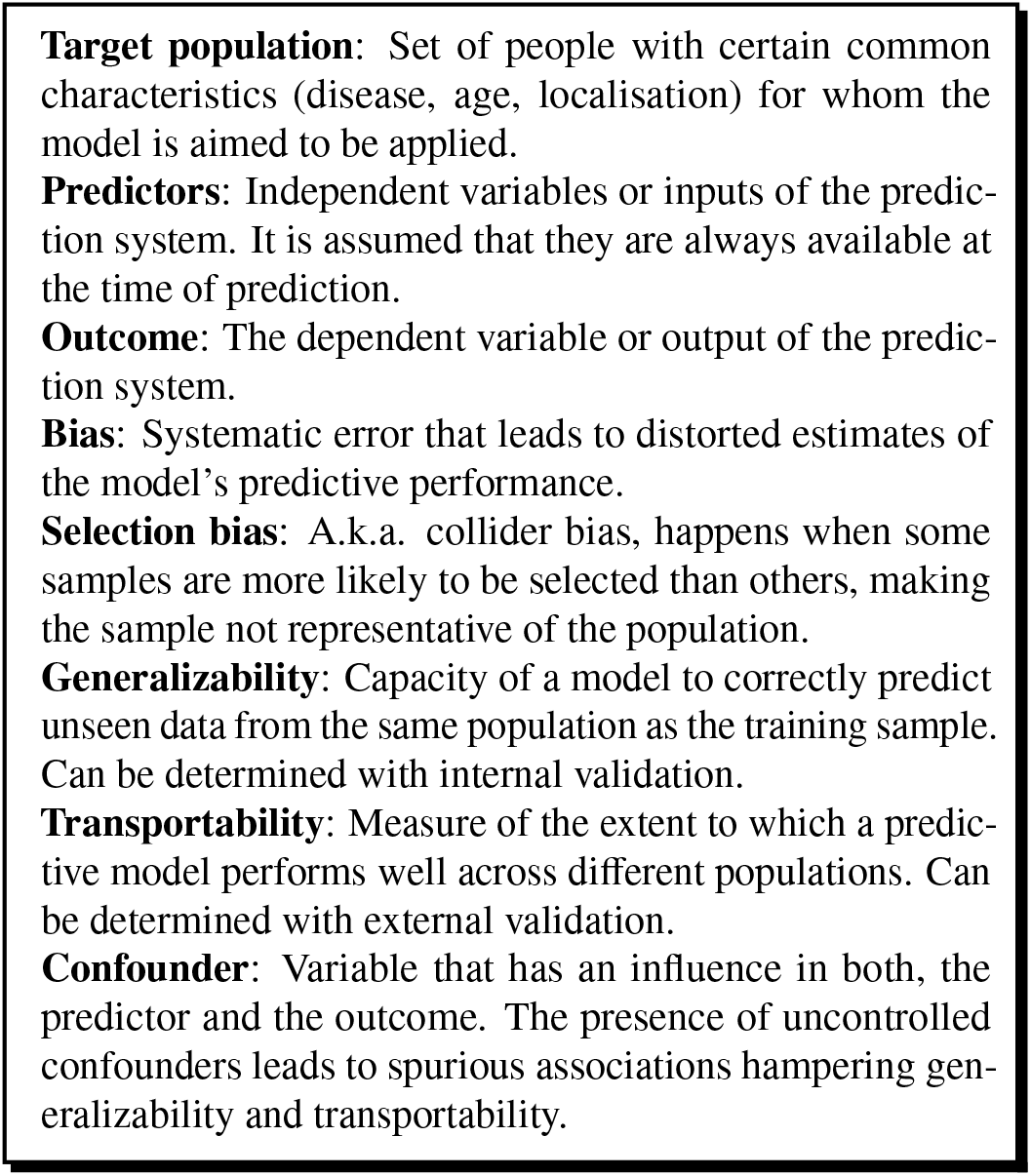
Key definitions

**Table 2.** Participant data. According to the available information, the data source type that best fits all the considered datasets is “cohort study”.

| Dataset | Eligibility & recruitment | Participant description | Treatment information | Study dates | Number of subjects/images | Country |
| --- | --- | --- | --- | --- | --- | --- |
| *Brixia*<sup>1</sup> | Yes<sup>6</sup> | Age, sex, location and study setting<sup>11</sup> | No | Yes | 2351 / 4703 | Italy |
| *ML Hannover*<sup>2</sup> | Unclear<sup>7</sup> | Sex | No | No | 71 / 243 | Germany |
| *CIA*<sup>3</sup> | Unclear<sup>8</sup> | Age, sex, race, location<br>weight, height and comorbidities | No | No | 105 / 256 | US |
| *HM Hospitales*<sup>4</sup> | Yes<sup>9</sup> | Age, sex and comorbidities | Yes | Yes | 2310<sup>12</sup> / 419 | Spain |
| *BIMCV-COVID19+*<sup>5</sup> | Yes<sup>10</sup> | Age and sex | No | Yes | 4706 / 16840 | Spain |
| *BIMCV-COVID19-* |  |  |  |  | 3238 / 6540 |  |

<sup>1</sup> *Brixia score COVID-19* dataset (Signoroni et al. (2020)) (https://brixia.github.io/).
<sup>2</sup> *COVID-19 Image Repository* (https://github.com/ml-workgroup/covid-19-image-repository).
<sup>3</sup> *Cancer Imaging Archive*, chest imaging with clinical and genomic correlates representing a rural COVID-19 positive population (COVID-19-AR) (https://wiki.cancerimagingarchive.net/pages/viewpage.action?pageId=70226443).
<sup>4</sup> *Covid Data Save Lives* (https://www.hmhopitales.com/coronavirus/covid-data-save-lives/english-version).
<sup>5</sup> *BIMCV-COVID19*, Valencian Region Medical ImageBank (Vayá et al. (2020)) (https://bimcv.cipf.es/bimcv-projects/bimcv-covid19/)
<sup>6</sup> All images from triage and patient monitoring in sub-intensive and intensive care units between March 4th and April 4th 2020 at the ASST Spedali Civili di Brescia.
<sup>7</sup> All data from the Institute for Diagnostic and Interventional Radiology, Hannover Medical School, Hannover, Germany.
<sup>8</sup> Patients who tested positive for COVID-19 from rural areas.
<sup>9</sup> Patients admitted with a diagnosis of COVID POSITIVE or COVID PENDING.
<sup>10</sup> All consecutive studies of patients with at least one positive PCR test or positive immunological tests for SARS-Cov-2. The the first iteration of the dataset was obtained from 11 hospitals in the Valencian Region (Spain) in the period of time between February 26th and April 18th, 2020.
<sup>11</sup> Sub-intensive and intensive care units. Age is given in 5 year intervals.
<sup>12</sup> From which only 189 subjects had accompanying X-ray images.

#### Potential outcomes

Outcomes were divided in two main categories, outcomes suitable for diagnostic models and for prognostic models, depending on whether the variable can be assessed at the time of image acquisition or if we must wait some time for the variable to change. A typical diagnostic outcome is the radiological report or annotation, which is available in three of the datasets, with different level of detail on each. Prognostic variables such as ICU admission and survival or discharge time were present in three datasets (see Table 3). It’s worth noting that the *Brixia* dataset contains severity diagnosis for some of the participants at multiple dates, and thus it can be used to build prognostic models of disease progression.

**Table 3.** Outcome information

| Dataset | Potential outcomes | Definition and methods | Blinded to predictors | Diagnostic or prognostic | Times<sup>1</sup> |
| --- | --- | --- | --- | --- | --- |
| *Brixia* | Brixia score<sup>2</sup> | Yes | No | Diag./Prog. | Yes |
| *ML Hannover* | ICU, survival | – | Yes | Prog. | Yes |
| *CIA* | ICU, survival | – | Yes | Prog. | Yes |
|  | Radiological findings | No | No | Diag. |  |
| *HM Hospitales* | ICU, survival, discharge | – | Probably not | Prog. | Yes |
| *BIMCV-COVID19±* | Radiological reports + findings | Yes | No | Diag. |  |
|  | PCR, IgG, IgM | Yes | Yes | Diag. |  |

<sup>1</sup> Time between predictor and outcome is particularly relevant for potential prognostic outcomes, and was reported only for these outcomes.
<sup>2</sup> Brixia score is a severity index of regional lung compromise (Signoroni et al. (2020)).

Regarding outcome definition and methods, lack of image annotations is not a critical issue because they can be assessed later by independent radiologists. However, if researchers are going to use the image annotations provided with the dataset, a precise definition and method description is needed, which are provided only for two datasets. Definition and method for ICU admission and survival were not considered necessary.

The only outcomes considered for diagnosis were features extracted from the images, and the image is necessary among the predictors. Therefore, for the diagnostic models considered in this work, the outcome can never be blinded to the predictor, and the predictor is always blinded to the outcome. As said previously, this could be useful for the analysis of the causal structure of the model. Regarding prognostic models, all predictors are blinded to future events determination unless a selection process is present, which can not be determined with the available data.

#### Candidate predictors

The amount of predictors, in addition to X-ray scans and demographic variables, vary widely between the datasets (see Table 4). Two datasets have no additional potential predictors, apart from image and demographics. All datasets include images in DICOM format, except for ML Hannover, that used NIfTI format for privacy reasons.

**Table 4.** Predictor information other than demographic variables.

| **Dataset** | **Candidate predictors <sup>1</sup>** | **Definition and methods** | **Timing** | **Blinded to outcomes** |
| --- | --- | --- | --- | --- |
| *Brixia* | modality, scanner manufacturer | Yes | Yes | Yes |
| *ML Hannover* | view, modality<br>laboratory data, vital signs | No | Yes | Yes |
| *CIA* | comorbidities | No | No | Yes |
| *HM Hospitales* | comorbidities, medications,<br>laboratory data, vital signs | Yes | Yes | Yes |
| *BIMCV-COVID19±* | modality | Yes | Yes | Yes |

<sup>1</sup> All datasets except *ML Hannover* included the DICOM headers, which contain further information about scanning setting, like view and scanner model, and could contain more information about the patient characteristics.

#### Sample size and missing data

Sample size is critical for Deep Learning prediction models, in particular when images are used as predictors, because of the high risk of overfitting due to the high dimensionality of the input. Lack of large enough sample sizes is a common issue in all medical applications, but COVID-19 data is specially scarce. Two of the reviewed datasets (*Brixia* and *BIMCV*) included a few thousand X-ray images, and the rest of them a few hundreds. See Table 2 for details.

Missing data was particular prominent in some potential predictors (laboratory data) of ML Hanover, however these variables are not widely used in image based Deep Learning models.

### 3.3. Frequently used COVID-19 datasets not included in the analysis

Several datasets that are frequently used in model building where deemed not eligible for the review, mostly because they are datasets aggregating primary sources (compilations) or they are collections of case studies. In the following, a brief overview is given. See to Fig. 3 for an overview of the composition and relation between these datasets and the primary sources of the data.

**Fig. 3.**
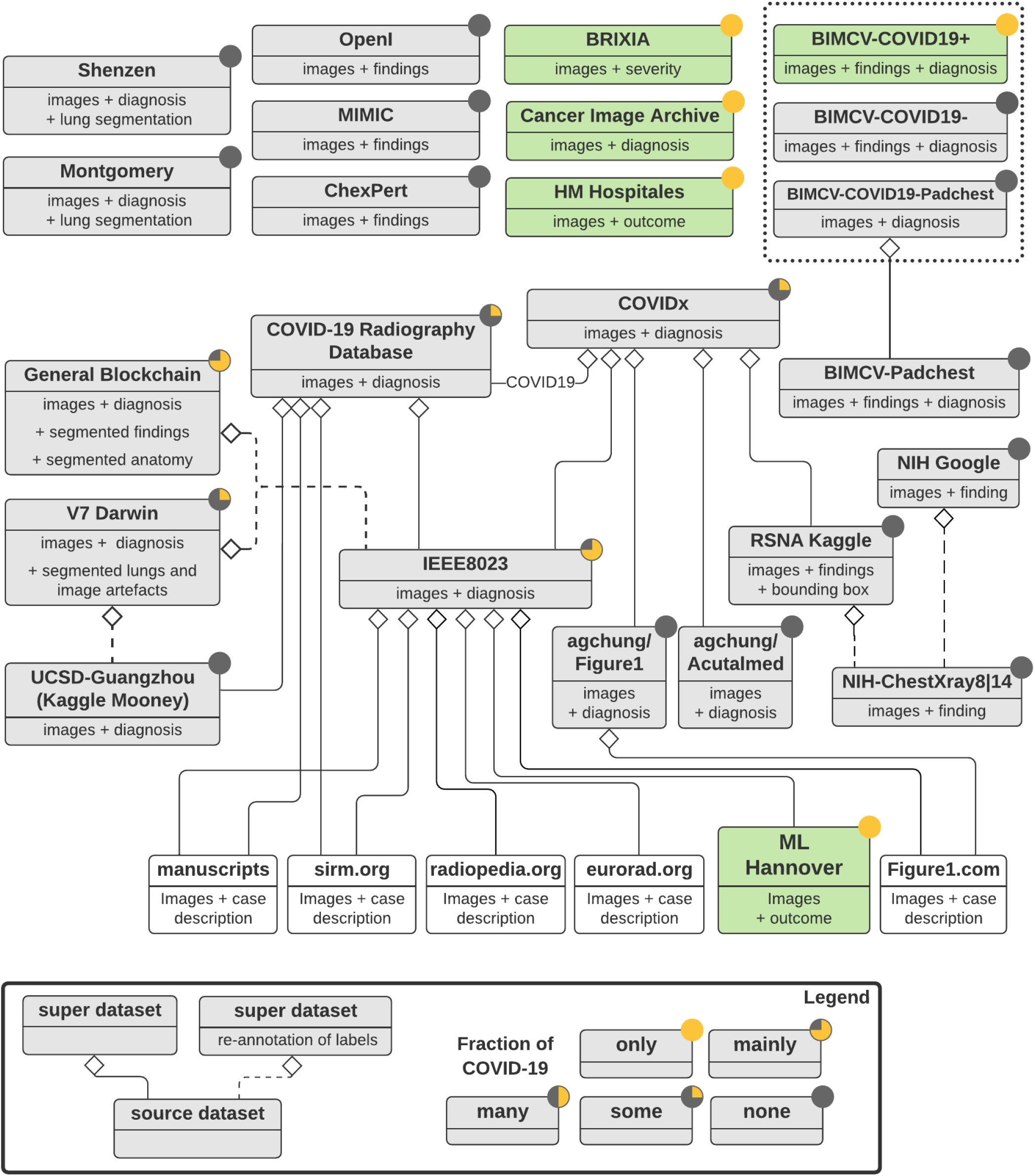
Overview of the relationships of popular COVID-19 and related non-COVID-19 datasets. Grey boxes represent datasets which can be downloaded as one entity. Green boxes indicate primary COVID-19 datasets meeting the inclusion criteria for review. Transparent boxes indicate data sources not directly available as downloadable datasets. Diamond shaped symbols indicate that the attached source dataset is (partially) included in an aggregated dataset. Dotted lines describe the case when images of a source dataset are included in an aggregated dataset but labels have been re-annotated. The colouring of the circle in the upper right corner of each datasets indicates the approximate proportion of COVID-19 patients (yellow) and control subjects (gray).

**Fig. 4.**
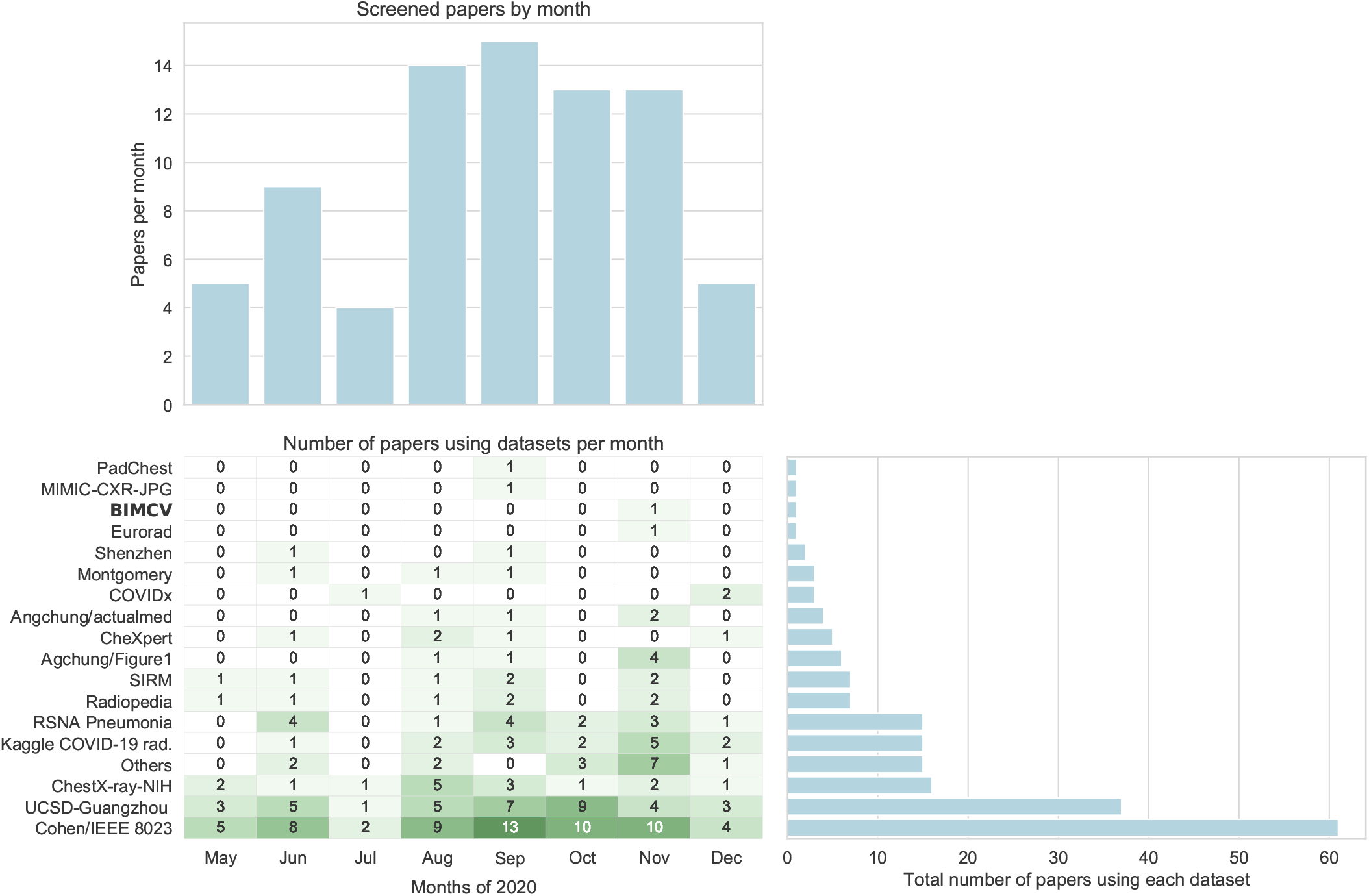
Temporal analysis of the datasets employed in the selected 78 peer-review papers. Top-left: Number of papers indexed in Pubmed that report a Machine Learning model for COVID-19 X-Ray imaging by month of their published date (from Pubmed database). Bottom-left: number of papers using a given dataset per month. Bottom-right: total number of papers using each dataset. Note that only one dataset among the classified as recommended were used (BIMCV), and only in one paper. The number of papers using exclusively Cohen/IEEE 8023 and UCSD-Guangzhou is 18, a particularly risky combination, as stated in Section 3.5

#### 3.3.1. Compilations and other datasets

Different radiological associations are making efforts to collect and provide images and reports on the internet to facilitate knowledge transfer of radiologist. These datasets are made public for educational reasons, and not for training prediction models. Among those, www.eurorad.org, www.sirm.org, www.radiopedia.org, www.figure1.com and www.bsti.org.uk provide images which are usually accompanied by a radiological description and varying depth of clinical information. Several initiatives such as Cohen/IEEE 8023 dataset collect and present this information in a structured dataset format.

The *Cohen*/*IEEE 8023* dataset (“COVID-19 Image Data Collection”, Cohen et al. (2020c)) is a collection of cases “extracted from online publications, websites, or directly from the PDF”, including the aforementioned case sharing websites. Additionally, it incorporates the ML Hannover dataset. It provides a well curated tabular view on the image meta-data containing the source document of the image, the imaging origin (hospital, city or country) and information about scanning view for most images, as well as gender, age and a variety of clinical features and outcomes for some cases. The dataset strongly focuses on different kinds of pneumonia (826 out 950 images) and therein especially on COVID-19 (584 out of 950 images) but provides very little cases of no finding (22). Additionally the dataset contains global severity scores for about 100 images created in a post-hoc analysis of images according to a severity scheme (Cohen et al. (2020a)). The Cohen/IEEE 8023 dataset was used to feed many other datasets. In the General Blockchain COVID-19 segmentations dataset two radiologist created, in a post-hoc annotation process, segmentation masks for anatomical parts, medical devices (e.g. tubes and probes) and radiological findings (Ground glass opacities and Consolidations).

The *agchung*/*Figure1* dataset contains images of 48 patients extracted from the case sharing website www.figure1.com. For some patients, it contains additional information about sex, temperature, pO2 saturation, the scanning view as well as some clinical notes. Most of images are assigned one of the labels COVID-19, pneumonia or no finding.

The *Kaggle COVID-19 radiography* database (Chowdhury et al. (2020)) aggregates COVID-19 cases from the Cohen/IEEE 8023, cases extracted from the sirm.org website and from 43 publications. Furthermore it incorporates control cases and cases of viral pneumonia from the UCSD-Guangzhou pediatric dataset. It does not include any meta-information, except the image dataset source.

The *V7 Darwin covid-19-chest-x-ray-dataset* also includes non-COVID-19 from the UCSD-Guangzhou pediatric dataset and COVID-19 cases from Cohen/IEEE 8023.

The *COVIDx* dataset (Linda Wang and Wong (2020)) is compiled from different sub-datasets. It contains the Cohen/IEEE 8023, the angchung/Actualmed and angchung/Figure1 datasets. It also includes COVID-19 cases from the COVID-19 radioagraphy database as well as Pneunomia and Normal cases from the RSNA Kaggle Pneumonia dataset. Each image is assigned one of the labels COVID-19, Pneunomia or Normal. As these sub-datasets contain common sources, there is the risk of case duplication due to incoherent source descriptions.

The *Actualmed* dataset contains images (CR, DX) of 215 patients. Each image is assigned a radiological diagnosis of “COVID19”, “inconclusive” or “No finding”. In addition it contains metadata of date and scanning view (AP vs PA). For the Actualmed dataset it was not possible to reliably determine the source (primary or secondary) of data due to lack of descriptive information.

### 3.4. Non-COVID-19 datasets

Some datasets built before the COVID-19 pandemic are frequently used for pre-training networks (Cohen et al. (2020a); Signoroni et al. (2020)) or to enrich training data with non-COVID-19 controls (Linda Wang and Wong (2020)). In general, these datasets are much larger and better curated than those for COVID-19.

The *Padchest* dataset (Bustos et al. (2020)) contains images (CR, DX, CT) of about 67000 patients together with radiological reports and thereof automatically extracted labels, “resulting in 22 differential diagnoses, 122 anatomic locations and 189 different radiological findings”. The dataset was collected from a single hospital (Hospital San Juan Hospital (Spain)) between 2009 and 2017 and contains DICOM meta-data. The aforementioned *BIMCV-COVID19-PADCHEST* is a subset of this dataset, reduced to samples of four classes (“Control”, “Infiltration without Pneumonia”, “Pneumonia without Infiltration” and “Pneumonia with Infiltration”).

*CheXpert* is a large dataset (Irvin et al. (2019)) released at the beginning of 2019 that includes 224,316 chest radiography from 65,240 patients collected from Stanford hospital. For fourteen classes of radiological findings a rule-based processing of the reports generated labels of presence, absence, or uncertainty thereof. Additionally this dataset comprises a dedicated testset, with 500 images annotated by consensus of eight radiologist.

The *ChestXray-NIH* dataset (Wang et al. (2017)) contains about 120k images of 30k patients from the clinical PACS database at National Institutes of Health Clinical Center. In its original version, all images were labelled with originally eight (ChestXray8) and in the current version fourteen (ChestXray14) different findings from thoracic pathology. The labels have been extracted from the corresponding radiological reports by Natural Language Processing. The underlying reports are not publicly available, but the dataset contains meta-information about gender, age and view position. Additionally there are annotated bounding boxes for all findings in about 1000 images.

The *RSNA Pneunomia* Kaggle dataset (Shih et al. (2019)) is a subset of 30k images from the ChestXray-NIH dataset with an enrichment of images with a Pneunomia related diagnosis. In a well defined annotation process a team of radiologist annotated areas of Lung Opacity with bounding boxes solely based on the information present in the image.

*ChestXray-NIH Google* is another subset of about 18k images from the ChestXray-NIH dataset with extra labels (Majkowska et al. (2020)). In another well designed annotation process radiologist assigned each image only with “access to the patient age and image view (PA/AP), but not to additional clinical or patient data” the presence/absence of four findings: pneumothorax, opacity, nodule/mass and fracture.

*Montgomery* dataset built in 2014 contains 138 frontal chest images collected in the department of health and human services, Mongomery Country, Maryland, USA. To each image, a short radiological report and a disease diagnosis is assigned (58 images with tuberculosis manifestations and 80 controls), as well as a lung segmentation annotation automatically generated under the supervision of a radiologist using anatomical landmarks (Candemir et al. (2013)). The images themselves contain written markings of the scanning view (AP and PA) and there is additional metadata about gender and age. The intended used of the dataset is to boost of computer-aided diagnosis for pulmonary diseases with focus on TB (Jaeger et al. (2014)).

The *Shenzhen* dataset was released together with the Montgomery dataset and contains images collected in collaboration the Shenzhen Hospital in China. It contains 662 frontal X-ray, of which 326 are normal and 336 contain TB manifestations. Additionally metadata includes sex, age and a short radiological description (Jaeger et al. (2014)).

*UCSD-Guangzhou* pediatric dataset contains more than 5000 chest X-ray images from *children* (AP view) selected from retrospective cohorts of pediatric patients of 1-5 year old from Guangzhou Women and Children’s Medical Center, Guangzhou (Kermany et al. (2018)). All images were assigned a diagnosis of viral/bacterial Pneumonia or Normal by two experts. No further meta-data is available.

*MIMIC-CXR-JPG v2*.*0*s.*0* is a large dataset comprising 377,110 chest x-rays associated with 227,827 de-identified imaging studies sourced from the Beth Israel Deaconess Medical Center. Images are provided with 14 labels derived from two natural language processing tools (NegBio and CheXpert) applied to the corresponding free-text radiology reports (Johnson et al. (2019)).

Indiana *University Chest X-rays*/*OpenI* is a dataset from the Indiana University created to provide a publicly available searchable database and comprises about 7500 Chest X-rays (Demner-Fushman et al. (2016)). It includes view information and radiological reports including main findings and impressions. In a well documented post-hoc annotation process the reports have been mapped to localised findings, e.g. “Cicatrix/lung/base/left”.

### 3.5. Analysis of dataset used on peer-reviewed papers

A total 151 papers indexed on Pudmed were identified. Next, 62 manuscripts were removed during the screening steps, due to: the datasets was not an X-ray modality (n = 49); it was an opinion paper, editorial, survey or review (n = 11); images were not from the lungs (n = 1); or the technology employed was not ML (n = 1). During the eligibility step, the 89 filtered papers were analysed to extract the datasets employed and all the paper that did not employ publicly available datasets (n = 8) or for which datasets could not be identified (n = 3) were removed. Finally, with the selected 78 papers, the datasets employed were extracted (Supplementary material, Appendix 3) and the temporal analysis was conducted.

Despite expecting a change in the pattern of dataset usage, as better resources became available, the contrary was observed, the pattern of dataset use of datasets was uniform. This may indicate that the dataset choices were made based on popularity of previously frequently used ones, even when better options had appeared. Shockingly, only one of those found during our analysis was used, and only in one article.

It’s also highlighted that 18 out of 78 papers employed exclusively the combination of “Cohen/IEEE 8023” and “UCSD - Guangzhou”. As we mention before, the combination of datasets can be dangerous, but this highly frequent combination is particularly dangerous because the “UCSD - Guangzhou” dataset was most probably employed for as a source of control cases, being children from 1 to 5 years old, while all COVID-19 came from adults, been this a clear source of confounding difficult to handle.

## 4. Discussion

This work highlights a common problem in imaging prediction models. While overfitting is commonly acknowledged when dealing with a small number of images, other sources of bias such as confounders and selection bias are not as frequently considered. This is evidenced by the careless use of datasets, where critical questions about populations, such as recruitment procedures, inclusion and exclusion criteria, or outcome measurement procedure, are not addressed. Some authors (Cohen et al. (2020c)) have already acknowledge that many datasets do not represent the real world distribution of cases, that the presence of selection bias is highly probable (particularly on case study collections), and therefore that clinical claims must take into account these limitations. However, the first step to tackle these issues is to have a good description of datasets in order to implement some strategy to reduce the bias. Or at least to be fully aware of model limitations and range of applicability. Unknown confounders and collider bias are not as problematic in prediction models as they are in causal inference (Griffith et al. (2020); Wynants et al. (2020)). However, model generalizability is compromised and its prediction power can only be maintained when training and target population remain similar and go through the same sampling mechanism. Even in this particular case, specifying the optimal target population cannot be done without knowing the training population characteristics. Recently, there are some recent efforts to address the general problem of bias in AI, and in particular regarding the use of human data. In Mitchell et al. (2019), for example, authors encourage transparent model reporting and propose a framework to describe many aspect of model building, including dataset description.

General considerations about clinical prediction model (Steyerberg (2009)) are as relevant in AI models as in linear regression models, although in the former case are much more difficult to address. Protocols for AI model development are being developed, in the meantime the minimum requirements for dataset description should be assessed (Collins and Moons (2019); Liu et al. (2019); Faes et al. (2020); Stevens et al. (2020)).

### 4.1. Bias in medical imaging ML

Medical Imaging Models, especially Convolutional Neural Networks (CNNs), are known not only to learn underlying diagnostic features, but also to exploit confounding image information. For example, it was shown that the acquisition site, regarding both the hospital system and the specific department within a hospital, can be predicted with very high accuracy (> 99%) (Zech et al. (2018)). If disease prevalence is associated with the acquisition site, as it is often the case, this can be a strong confounder. Thus, in any composite dataset having separated sub-datasets for COVID-19 and control cases, the dataset is completely confounded with the group label and therefore it is difficult to isolate the disease effect from dataset effect, making learning almost impossible and posing a high risk of overestimating prediction performance. Indeed it has been observed that, by training on different COVID-19 and non-COVID-19 dataset combinations, the “deep model specialises not in recognising COVID features, but in learning the common features [of the specific datasets]” (Tartaglione et al. (2020)). Eventually, a CNN model is able to identify the source dataset with a high accuracy (> 90%) solely from the image border region containing no pathology information at all (Maguolo and Nanni (2020)).

Besides acquisition site, the demographic characteristics of populations can also be a strong confounder. Datasets that take cases from the UCSD-Guangzhou pediatric dataset as non-COVID-19 examples (maximum age 5 years old) pose the risk that models will associate anatomical features of age with the diagnosis since, for example in the Cohen/IEEE 8023 dataset, the minimum age is 20 years old (mean 54). Controlling for confounders is already difficult in deep learning models, and normalising the images in such a wide and disjoint range of ages seems an impossible task.

But one not only has to be wary in composite datasets, also single source datasets are not free of potential confounders and other sources of bias. The classical example would be a different imaging protocol depending on the patient’s health status. For example, the PA erect view is the preferred imaging view in general, but if the patient is not able to leave the bed it is much more common to do an AP view image.

Another confounding factor might be the presence of medical devices like ventilation equipment or ECG cables, which allows a model to associate images with patient treatment instead of disease status. For example, for the NIH ChestXray14 dataset, a critical evaluation has shown that “in the pneumothorax class, […] 80% of the positive cases have chest drains. In these examples, there were often no other features of pneumothorax” (Oakden-Rayner (2020)). Datasets which provide additional annotations on the presence of medical devices (e.g. *BIMCV, 7labs, General Blockchain*) facilitate a risk analysis on this confounding effect and also enable mitigation strategies in training.

In general, one has to distinguish between labels which have been annotated by taking only the image itself into account, and labels which have been generated by a different source, i.e. from another diagnostic method like CT or PCR. Unfortunately, radilogical reports done in a in clinical routine are a mixture of both. Radiologist are often aware of the patients clinical context and this information is reflected in the reports, since they are done with the aim of communicating information between different doctors. For example, it has been shown for the NIH ChestXray14 dataset that, in a substantial fraction of images, the associated finding extracted from the reports can not be confirmed by a post-hoc assessment of the the images (Oakden-Rayner (2020)).

Biases arise more easily when outcome labels and prediction model intended application is not clearly defined. If the model objective is to find radiological manifestation of the disease in the images that are not necessarily apparent to the radiologist naked eye, the labels should be the best possible diagnostic assessment obtained by any diagnostic test that doesn’t include image information from same modality. For example, a perfectly righteous goal could be to determine whether a feature observed in CT, but not visible in XR, could be detected by subtle signals that ML models can identify. In contrast, if the goal is to reproduce radiological findings (for example, to save radiologist time) the label should be radiological annotations assessed by an independent clinician that has no information except for the image. Otherwise the risk of bias increases sensibly and the generalisation ability is compromised, because we can not really understand where the key information is coming from, what the model is learning, and what the possible sources of bias are. In this sense, it’s worth noting that a couple of datasets do provide such annotations solely derived from the images (RSNA Kaggle, NIH Google, General Blockchain, 7labs).

### 4.2. Advice for modellers

To avoid risk of bias coming from dataset misuse is is important that researchers follow transparent practices and adequate reporting guidelines. For reviewers to assess whether the chosen datasets are appropriate for the research question or intended use, it is necessary that researchers address the following:

- Merging subjects from different datasets should be done from original sources only, so that potential bias can be easily evaluated from the study and is not hidden in the data source. The selected images from each dataset should be listed one by one (e.g. as supplementary material), including all the patients available information. The reasons behind the inclusion of subjects from these particular datasets and with these specific characteristics should be explained in the context of the intended use of the model.
- The strategy followed to mitigate the potential biases should be explained. For example, datasets could be re-balanced or re-weighted (Jiang and Nachum (2019); Amini et al. (2019)) in terms of outcome prevalence for each of the key demographic variables.
- Ask oneself which population is represented by the datasets, i.e. which were the recruitment procedures, location and setting, the inclusion and exclusion criteria, and subjects demographics. They should also address how exactly the outcome was obtained and how is related with the disease and with the application.
- Explain how the model can be applied to a clinical setting, which is the benefit for the patient or how it would help medical personal to make decision.

## 5. Conclusion

This work present a first attempt to systematically evaluate imaging datasets in terms of their utility to train predictions models. We follow PRISMA guidelines to systematically search for X-ray chest images databases of COVID-19 subjects, either screening papers reporting models where these images are used or directly searching for datasets in dataset search engines and compilations of datasets. Inspired by PROBAST, TRIPOD and TREE statements, this work aimed to answer whether the available COVID-19 X-ray datasets could be used to train or validate clinical prediction models with low risk of bias. With this objective in mind, the CHARMS checklist was adapted to extract the relevant information about participants, outcomes, predictors and sample size.

The information provided in all the reviewed datasets is too scarce to guaranty that a model can be built with low risk of bias. For example, key questions regarding participant information and their appropriateness for a given application can not be answered. This finding is consistent with results presented in a systematic methodological review of Machine learning for COVID-19 prediction models using chest X-rays and CT scans (Roberts et al. (2021)), where PROBAST assessment rated all X-ray-based models as having a high or unclear risk of bias in the Participant domain.

Larger datasets that include better study design and more comprehensive descriptions are being built and are becoming available to researchers (as a data collection). However, they are provided under much stringent conditions and a larger project evaluation process must be undergone. New and old models should be developed and tested using these datasets. Up to our knowledge, there are currently no prediction models reported that have used these datasets.

It is urgently needed that more images from larger and better datasets are made publicly available. Dataset owners should make an effort to improve documentation about whole dataset building process to increase significantly the dataset value and the quality of models trained on them. For example: there should be a clear statement of dataset intended use, and explicit warning of common misuse cases; label definition and generating procedure should be reported in detail, so that other researchers can verify accuracy of label assignments and evaluate the utility and adequacy to the problem at hand; finally, datasets should contain cohort characteristics and subject selection criteria information, in order to evaluate the risk of selection bias and to check if the training and target population has similar characteristic.

Contrary to classical statistical models or standard machine learning methods, deep learning models are highly complex systems that may have several building steps. For reasons that range from avoiding overfitting to reducing memory and computation needs, some parts of the models are sometimes pre-trained, using specific dataset, and then kept fixed as other parts of the model are finetuned later. Quality standards of datasets used to pretrain these building blocks may not necessarily be as high as the ones for finetuning the final model, and some of these datasets that are deemed close to useless for training a serious medical diagnostic tool may be perfectly appropriate for pre-trainig steps. One only has to be cautious not including those subjects in the cross-validation loops of the final model.

Current open access datasets could be useful for pre-training models, or to illustrate model features and potential applications. However, for models trained exclusively with these open datasets, claims about efficacy could be highly biased, and generalizability and transportability are uncertain. Applicability to clinical settings is therefore extremely risky and not recommended.

All in all, both *Brixia* and *BIMCV* provide a next level of dataset quality and quantity for diagnostic models compared to open datasets abailable in the beginning of the pandemic and can serve as new grounds for modellers. From the reviewed datasets, these two were the only ones with a sample size large enough (thousand of images) to start training a DL model, while the others have only a few hundreds. Furthermore, these were the only two datasets among all the identified datasets (not only the reviewed) that provided a description of the inclusion and exclusion criteria, despite of falling short to fully describe the population they are representing. Unfortunately, neither of the two provided accompanying predictors (such as laboratory data or other clinical reports), treatment or comorbidity information, which was sometimes present in some of the other datasest.

This review should help modellers to choose the appropriate dataset for their modelling needs and to raise awareness on biases to look out while training models. It is also encouraged that everyone validates their models and reports benchmarking results on a possibly small but very well curated external dataset, which is carefully selected to represent the real clinical use case as close as possible.

Although dataset quality is the most important requirement for a medical diagnostic system to be reliable, other aspects of the model building are also prone to biases. Adherence to transparent practices, such as the TRIPOD reporting guideline or TREE critical questions, and assessing risk of bias with PROBAST tool, would be a starting point (Parikh et al. (2019); Sounderajah et al. (2020)). However extensions of these guidelines are required in order to be fully applicable to deep learning systems (Wynants et al. (2020)). Many efforts are already being done in this direction: extension of TRIPOD and CONSORT/SPIRIT (TRIPOD-ML, Collins and Moons (2019)) and CONSORT-AI/SPIRIT-AI (Liu et al. (2019)) statements are being developed, focused on model validation and clinical trials, respectively; considerations for critically appraising ML studies are given in Faes et al. (2020); and reporting recommendations are given in Stevens et al. (2020).

## Data Availability

All data used is publicly available

## Acknowledgments

This work was supported by the Luxembourg National Research Fund (FNR) COVID-19/2020-1/14702831/AICovIX/Husch grant. Beatriz Garcia Santa Cruz is supported by the FNR within the PARK-QC DTU (PRIDE17/12244779/PARK-QC). Andreas Husch is partially supported by the Fondation Cancer Luxembourg. The authors would like to thank Prof. Peter Gemmar, Trier University of Applied Sciences, Trier, Germany and Prof. Frank Hertel, Centre Hospitalier de Luxembourg, Luxembourg, for discussions.

## Footnotes

1 https://datasetsearch.research.google.com

2 http://open-source-covid-19.weileizeng.com/tags

3 https://github.com/HzFu/COVID19_imaging_AI_paper_list#dataset

4 https://aimi.stanford.edu/resources/covid19

