## Supplementary_Material_1 for "Public Covid-19 X-ray datasets and their impact on model bias - a systematic review of a significant problem"

### Supplementary Material, Appendix 1.

Table S1: Adapted items from CHARMS tool.

| **Domain** | **Key items** |
|---|---|
| Source of data | cohort, case-control or cross-sectional |
| Participants | Participant eligibility and recruitment method |
| | Participant description |
| | Details of treatments received |
| | Study dates |
| Outcomes | List of potential outcomes |
| | Definition and method for measurement of outcome |
| | Was the outcome assessed without knowledge of the candidate predictors (i.e., blinded)? |
| | The outcome is useful for a prognostic or a diagnostic models? |
| | Time of outcome occurrence or summary of duration of follow-up |
| Candidate predictors | List of candidate predictors |
| | Definition and method for measurement of candidate predictors |
| | Timing of predictor measurement |
| | Were predictors assessed blinded for outcome, and for each other? |
| Sample size | Number of participants and number of outcomes/events |
| Missing data | Number of participants with any missing value |
| | Number of participants with missing data for each predictor |
