## Supplementary_Material_2 for "Public Covid-19 X-ray datasets and their impact on model bias - a systematic review of a significant problem"

### Supplemental Material, Appendix 2.

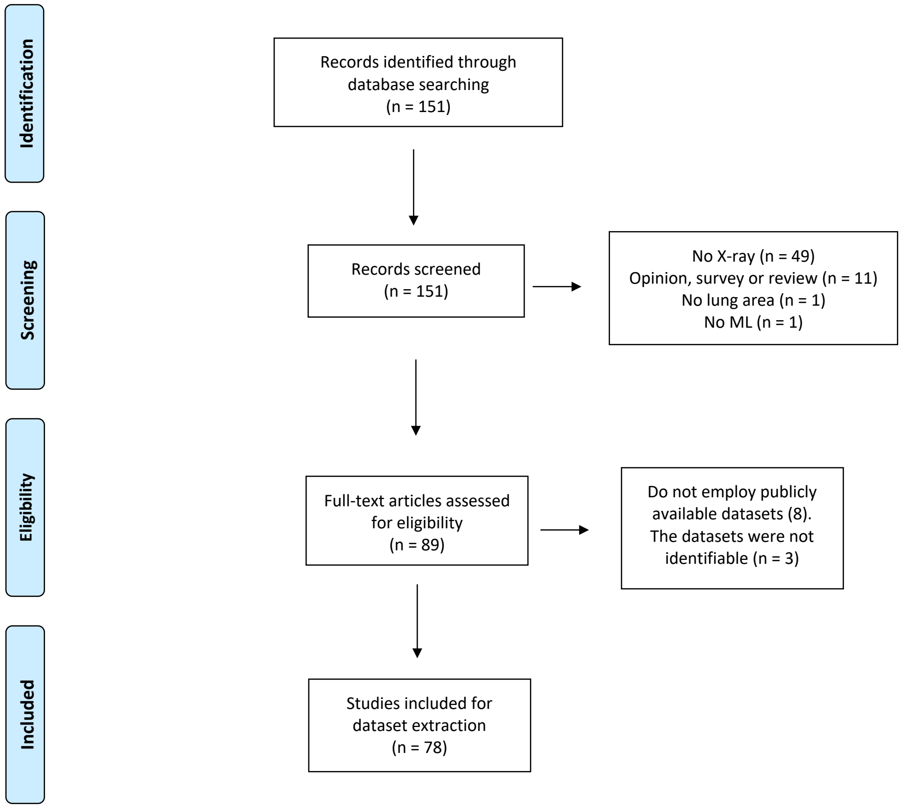


Figure S1: PRISMA workflow for analysis of the dataset usage frequency.
